# Employee Performance Appraisal Methods: A Scoping Review

**DOI:** 10.1101/2025.07.02.25330701

**Authors:** Mobarakeh Alipanah Dolatabad, Pedram Nourizadeh Tehrani, Hamid Pourasghari, Nasrin Sharfafchizadeh

## Abstract

**Background:** Employee performance appraisal is essential for improving healthcare service delivery through systematic staff evaluation. In the health sector, effective appraisals support decision-making and professional development.

**Objective:** This study aims to map and categorize existing employee performance appraisal methods in the health system using a scoping review approach.

**Methods:** This scoping review was conducted using the JBI 2024 protocol. Articles published up to December 2024 were identified through four databases: PubMed, Scopus, Web of Science, and Google Scholar. The keywords used were “Personnel appraisal,” “Health workers,” and “Health workforce.” Inclusion criteria focused on studies assessing employee appraisal methods in health systems.

**Results:** From 1245 initially identified articles, 42 met the inclusion criteria. Appraisal methods were classified into traditional and modern categories. Traditional methods included ranking, critical incidents, and graphic rating scales. Modern approaches involved 360-degree feedback, Management by Objectives (MBO), and Behaviorally Anchored Rating Scales (BARS).

**Conclusion:** No single appraisal method suits all healthcare environments. A hybrid approach tailored to organizational context and job roles is recommended. Emphasis should be placed on objective evaluation, customization, and rater relevance to improve performance outcomes.

## Introduction

Performance appraisal (PA) systems are fundamental tools within human resource management (HRM) that systematically assess employee performance and contributions toward organizational goals. These systems serve multiple purposes, including providing feedback, guiding employee development, supporting administrative decisions such as promotions and compensations, and enhancing overall organizational effectiveness [1,3,11]. The concept of PA has evolved from simple annual evaluations to multifaceted processes integrating continuous feedback, goal setting, and performance coaching [4,5]. Effective PA contributes significantly to employee motivation and job satisfaction, which in turn impact organizational commitment and productivity [6,7,13].

Despite their importance, many organizations struggle with implementing effective PA systems. Common challenges include evaluator biases, lack of clarity in performance criteria, insufficient training of raters, and poor communication between managers and employees [14,15]. These challenges often lead to employee dissatisfaction and decreased trust in the appraisal process, undermining its intended benefits [16]. Furthermore, PA systems that do not account for contextual factors, such as organizational culture or external pressures, may fail to achieve their goals [12].

In sectors like healthcare and public administration, where employee performance directly affects service quality and public welfare, the stakes are even higher. Healthcare professionals face unique challenges in PA, such as evaluating qualitative outcomes and balancing clinical duties with administrative responsibilities [14,16]. In recent years, technological advancements and the rise of remote work—accelerated by the COVID-19 pandemic—have further complicated traditional PA methods, demanding innovative approaches to appraisal [17]. The pandemic has forced organizations to adapt by implementing virtual assessments, increasing reliance on self-appraisals, and emphasizing more frequent and flexible feedback cycles [17].

Moreover, the theoretical foundations of PA draw heavily on motivational theories such as Self-Determination Theory, which emphasizes the importance of autonomy, competence, and relatedness in fostering employee engagement and performance [8]. Aligning PA practices with these psychological needs through participatory approaches like Management by Objectives (MBO) can enhance employee satisfaction and organizational outcomes [9]. Empirical evidence suggests that employees who perceive PA systems as fair and developmental report higher motivation and commitment, whereas perceptions of bias and unfairness lead to disengagement and turnover intentions [13,15].

Given the wide range of PA methods—ranging from traditional rating scales to multifactorial evaluation models and 360-degree feedback—there is a need to synthesize current evidence to guide practitioners and researchers [10,12]. This scoping review seeks to map the landscape of PA research, identify common appraisal methods, explore their impacts on employee satisfaction and organizational effectiveness, and highlight barriers and facilitators for successful implementation. Additionally, the review addresses adaptations in PA systems in response to emerging challenges such as global health crises.

By providing a comprehensive overview of PA systems, this review aims to support HR professionals, policymakers, and organizational leaders in designing appraisal processes that are evidence-based, contextually appropriate, and capable of driving continuous improvement at both individual and organizational levels.

## Materials and Methods

### Study Design

This scoping review follows the methodological framework outlined by JBI 2024. Scoping reviews are particularly suited for mapping broad topics, identifying key concepts, gaps in research, and types of evidence available, without restricting to quality appraisal as systematic reviews do. This approach allows for a comprehensive understanding of performance appraisal (PA) systems across diverse organizational settings and contexts.

### Search Strategy

A systematic search was conducted across multiple electronic databases, including PubMed, Scopus, Web of Science, and Google Scholar, to capture a wide range of literature published up to April 2025. The search terms combined keywords related to performance appraisal, employee satisfaction, organizational outcomes, and appraisal methods. Boolean operators were used to refine the search: (“performance appraisal” OR “performance evaluation” OR “employee evaluation”) AND (“employee satisfaction” OR “job satisfaction”) AND (“organizational performance” OR “organizational commitment”). Additional hand searches were performed on reference lists of key articles and relevant reviews.

### Inclusion and Exclusion Criteria

Studies were included if they:

- Focused on performance appraisal systems or methods in organizational settings.
- Reported outcomes related to employee satisfaction, motivation, or organizational performance.
- Were empirical studies (quantitative, qualitative, or mixed methods), reviews, or theoretical papers.
- Published in English.

Studies were excluded if they:

- Did not address PA systems or employee-related outcomes.
- Were opinion pieces without empirical data.
- Were conference abstracts without full text available.

### Data Extraction

Two independent reviewers screened titles and abstracts for eligibility, followed by full-text review to confirm inclusion. Discrepancies were resolved by discussion or consultation with a third reviewer. Extracted data included study characteristics (author, year, country, setting), PA methods examined, main findings on employee and organizational outcomes, and identified challenges and facilitators in PA implementation.

### Data Synthesis

A narrative synthesis approach was employed to summarize findings, organized by themes such as appraisal methods, effects on employee satisfaction, motivational aspects, and contextual factors influencing PA effectiveness. Quantitative results were tabulated where appropriate to illustrate trends and gaps in the literature.

## Results

### Study Selection

The initial database search yielded 1,245 articles. After removing duplicates, 1,030 titles and abstracts were screened. Of these, 215 articles were selected for full-text review based on inclusion criteria. Finally, 42 studies were included in the scoping review for detailed analysis (table 1).

**Table 1.** Summary of Performance Appraisal Methods and Their Characteristics.

| Method | Description | Advantages | Disadvantages | References |
| --- | --- | --- | --- | --- |
| 360-Degree Feedback | Collects performance data from supervisors, peers, subordinates, and sometimes customers. | Comprehensive feedback, promotes self-awareness | Time-consuming, potential bias, requires trust | 3, 13, 15 |
| Management by Objectives (MBO) | Employees and managers set specific measurable goals together and review progress periodically. | Goal clarity, motivation enhancement | May focus too much on goals, neglect other factors | 4, 9 |
| Rating Scales | Uses a numerical scale to evaluate various performance factors. | Simple and easy to use | Subjective bias, may not capture complexity | 3, 10, 12 |
| Behaviorally Anchored Rating Scales (BARS) | Combines qualitative and quantitative assessments based on specific behaviors. | More objective, behavior-focused | Complex to develop, time-consuming | 5, 11 |
| Checklist Method | Evaluator checks statements that describe employee behaviors. | Quick and easy | Lacks depth, may not reflect actual performance | 3, 16 |
| Essay Method | Evaluator writes a detailed narrative about employee's performance. | Provides rich information | Time-consuming, subjective | 3, 16 |
| Forced Distribution | Employees are ranked and placed in performance categories (e.g., top 10%, average 70%). | Identifies top and low performers | Can demotivate employees, competition fostered | 3 |

### Characteristics of Included Studies

The included studies spanned a variety of industries and organizational settings, including healthcare, telecommunications, finance, manufacturing, and public services. Most studies were conducted in Asia (35%), Africa (25%), Europe (20%), and North America (15%). The publication years ranged from 2005 to 2025, with a notable increase in research on PA systems in the last decade.

### Performance Appraisal Methods Identified

The studies described a diverse range of PA methods:

- Traditional rating scales and ranking systems [12,15]
- Management by Objectives (MBO) [9]
- 360-degree feedback [5,13]
- Multifactorial evaluation models [12]
- Self-assessment combined with supervisor evaluation [7,16]

The selection of methods was often influenced by organizational culture, size, and technological infrastructure.

### Impact on Employee Satisfaction and Motivation

A significant number of studies reported a positive correlation between well-implemented PA systems and employee satisfaction [2,6,7,13]. Performance appraisals that included clear goal-setting, constructive feedback, and opportunities for employee participation tended to enhance motivation and job commitment [8,9]. Conversely, studies highlighted dissatisfaction arising from perceived bias, lack of transparency, and inadequate training of appraisers [14,16].

### Organizational Outcomes

Effective PA systems were linked to improved organizational performance metrics such as productivity, reduced turnover, and enhanced service quality [3,10,11]. Several studies emphasized the role of PA in aligning individual objectives with organizational goals, thus fostering a performance-oriented culture [3,11].

### Challenges and Barriers

Common barriers to effective PA implementation included:

- Inconsistent application of appraisal criteria [14,16]
- Lack of managerial skills in conducting appraisals [16,17]
- Resistance from employees due to fear of negative evaluations [15]
- Technical limitations and insufficient data management systems [12]

### Facilitators for Successful Performance Appraisal

Studies highlighted the importance of:

- Training for managers and employees on PA processes [5,16]
- Incorporation of technology to streamline appraisal and feedback [12]
- Transparent communication and involvement of employees in appraisal design [6,13]
- Regular review and updating of appraisal criteria to reflect changing job roles [10]

## Discussion

This scoping review synthesized evidence on performance appraisal (PA) systems, highlighting their diversity in methods, impacts, challenges, and facilitators across multiple sectors and regions. The findings confirm that while PA systems are widely recognized as crucial tools for managing employee performance and fostering organizational growth, their effectiveness heavily depends on implementation quality and contextual factors.

Consistent with previous research [3,7,13], our review showed that PA methods such as Management by Objectives (MBO) and 360-degree feedback can enhance employee motivation and satisfaction when applied with transparency and fairness. This aligns with Self-Determination Theory [8], emphasizing the role of autonomy, competence, and relatedness in motivating employees. The involvement of employees in the appraisal process and clear communication of goals emerged as key factors contributing to positive outcomes.

However, the review also revealed pervasive challenges including appraisal bias, inadequate training of evaluators, and resistance from employees, echoing concerns raised in studies by Giangreco et al. [14] and Nikpeyma et al. [16]. These barriers often undermine the credibility of PA systems and reduce their impact on organizational performance. Notably, technological limitations remain a critical issue, suggesting a need for investment in digital solutions that can support objective data collection and analysis [12].

The review underscores the importance of ongoing training and capacity building for managers, as well as the need for periodic review and adaptation of appraisal criteria to remain aligned with evolving job demands [5,10,16]. Organizations that foster a culture of continuous feedback and development rather than punitive assessment are more likely to realize the full benefits of PA systems [13,15].

## Conclusion

Performance appraisal systems are essential mechanisms for aligning individual performance with organizational goals, enhancing employee satisfaction, and driving organizational effectiveness. However, successful implementation requires careful design, transparent communication, evaluator training, and adaptation to organizational context. Future research should explore innovative appraisal models leveraging technology and focus on longitudinal outcomes to better understand the sustainability of PA system benefits.

## Data Availability

N/A

## Authors contributions

**Supervision:** Hamid Pourasghari, Nasrin Sharbafchizadeh

**Methodology:** Nasrin Sharbafchizadeh

**Data collection:** Mobarakeh Alipanah, Pedram Nourizadeh Tehrani

**Writing – original draft:** Mobarakeh Alipanah, Pedram Nourizadeh Tehrani

**Writing – review & editing:** Nasrin Sharbafchizadeh

## Notes

### Competing Interest Statement

The authors have declared no competing interest.

### Funding Statement

The author(s) received no specific funding for this work.

